# Spatiotemporal patterns of African infant hydrocephalus are predicted by prenatal environment and ancestral genomics

**DOI:** 10.64898/2026.03.10.26347974

**Authors:** Lucinda Newbury, Ronald Mulondo, Misaki Sasanami, Mercedeh Movassagh, Brian Kaaya Nsubuga, Davis Natukwatsa, Joshua Magombe, Esther Nalule, Moses Wabukoma, Paddy Ssentongo, Joseph N. Paulson, Edith Mbabazi-Kabachelor, Benjamin C. Warf, Peter Ssenyonga, Justin Onen, Jessica E. Ericson, Julius Kiwanuka, Sarah U. Morton, Peter J Diggle, Andrew J. Whalen, Claudio Fronterre, Steven J. Schiff

## Abstract

Infant hydrocephalus is the leading indication for neurosurgery in early childhood in sub-Saharan Africa. The causes and consequences of hydrocephalus generate a high rate of morbidity and mortality. Postinfectious hydrocephalus and neural tube defects together comprise the majority of infant hydrocephalus in such settings, and both conditions are in principle preventable. In this analysis of data including more than 5000 individuals treated for infant hydrocephalus over 19 years in Uganda, we find strong evidence that prenatal environmental exposures predict the risk of hydrocephalus. The risk of postinfectious hydrocephalus was positively correlated with poverty and local rainfall (with maximal rainfall effect 14 days prior to birth), and negatively correlated with geographical ancestral admixture and elevation. The risk of neural tube defects was positively correlated with geographical ancestral admixture and negatively correlated with vegetation index (with maximal vegetation effect 8 months prior to birth). These findings associating potential prenatal gene-environment interactions with infant hydrocephalus lay the foundation for more targeted and optimized public health policies to reduce the rates of these devastating conditions.

## 1 Introduction

Hydrocephalus in infants is the leading indication for neurosurgery in early childhood in sub-Saharan Africa (*1*). In Uganda almost 60% of infant hydrocephalus is acquired as a consequence of a severe infection following birth, such as neonatal sepsis. (*2*). In recent years, a single species of bacteria has been found responsible for the majority of postinfectious hydrocephalus (PIH) in Uganda (*3–5*). Such acquired PIH differs from congenital causes of hydrocephalus (*6*). The most common congenital causes of hydrocephalus in low- and middle-income countries are neural tube defects (NTD) such as spina bifida (*1*). In infants with spina bifida, hydrocephalus commonly develops following surgical closure of an open myelomeningocele (MM) (*7*).

In principle, both PIH and NTD are preventable, with more effective prevention or treatment of infection in the case of PIH (*5*), and by micronutrient supplementation prior to conception and fetal closure of the neural tube in early pregnancy in the case of MM (*8*). Optimal prevention of these conditions requires an accurate delineation of the geographical and temporal patterns of highest incidence and the targeting of resources for maximal risk reduction.

There is growing evidence of geographical and seasonal patterns in the distribution of hydrocephalus cases. In (*9*), it was shown that PIH follows a pattern related to the biannual rainy seasons of the East African Highlands (*10*). More recently, *Paenibacillus thiaminolyticus*, which also has a distinct geographical distribution, was discovered to cause a unique syndrome, neonatal paenibacilliosis (*5*), that leads to an extraordinarily high rate of PIH in survivors (*4*). No evidence of maternal vertical transmission has been observed for such *Paenibacillus* infections (*11*), but subsequent neonatal infections can occur within the first week following birth (*5*), consistent with an environmental source. It is increasingly recognized that genetic variation can be associated with the risk of infection (*12–14*). Recently, prenatal environmental effects have been linked to subsequent childhood growth failure (*15*), with concerns for both prenatal nutritional and infection risk (*16–18*).

The open spinal defect MM is a survivable form of NTD. The malformation occurs early in pregnancy (about the 28th day of gestation), and its risk is well known to be increased when the maternal diet is deficient in folate (vitamin B9) (*19*). Such congenital malformations can also have a strong genetic basis; indeed, parents having had one child with MM have a 10-100 fold increased risk of a second child with this malformation. Nevertheless, the human genetic underpinnings of this effect have not been well characterized. Population studies have demonstrated that single gene variations can substantially affect maternal blood levels of folate following dietary supplementation, and that these genomic variations vary widely geographically (*20, 21*).

Congenital forms of hydrocephalus that are unrelated to NTDs (which we refer to as non-postinfectious hydrocephalus, NPIH) constitute a variety of phenotypes. Because each phenotype has varying risk factors, their combined average incidence is distributed more evenly throughout the population (*22*). In particular, the incidence of NPIH is more uniform than that of PIH (*4*) or NTD, making it valuable as a case control contrast. This is especially true for PIH, since the clinical phenotype in early childhood – macrocephaly – is identical to that of NPIH, which means that referral bias will not be introduced as a consequence of differing clinical presentations.

In previous work (*23*), we fit a generalized linear mixed effects model to explore the associations between environmental factors and the relative risk of MM. In that work, an increase in the normalized difference vegetation index (NDVI) appeared to significantly reduce risk. It is possible to extend this investigation by using the enhanced vegetation index (EVI), an alternative vegetation metric (*24*). The EVI improves upon the NDVI by reducing saturation in high-biomass areas and correcting for atmospheric and soil background effects, thereby providing more reliable measurements (*25, 26*). In addition, there appeared to be spatial factors that were difficult to account for without access to the ancestral genomics analysis that we now have. Because MM risk is known to have both a genetic component and a nutritionally modifiable effect on risk, those preliminary findings motivated the more exhaustive and detailed examination of the potential gene-environment interactions of this present report.

We here characterize the spatiotemporal distribution of the relative risks of PIH and MM by analyzing thousands of cases of Ugandan infant hydrocephalus over 19 years as a function of genomic ancestry and environmental correlates. We characterize unexpected gene-environment risk factors (*27*) with important implications for prediction and prevention of these diseases.

## 2 Methods

### 2.1 Data

#### 2.1.1 Ethics Statement

To study the genomes of the Ugandan cohorts, ethical oversight was provided by the Mbarara University of Science and Technology Research Ethics Committee, the CURE Children’s Hospital of Uganda Institutional Review Board, the Pennsylvania State University and Yale University Institutional Review Boards, and the Ugandan National Council on Science and Technology. The participants were recruited as previously described as part of studies of infant hydrocephalus and neonatal sepsis in Uganda (*3–5*) and samples were collected prior to the current Genome Data Sharing policy by the US National Institutes of Health. Specimens were shared with US institutions under Material Transfer Agreements and US Centers for Disease Control permits. A Genome Data Sharing Policy was screened by NIH and approved by all the above ethics and government oversight committees which, in brief, allows for the results of genomic analysis to be shared only in aggregate, so that individual participants cannot be re-identified. The data sharing agreement also stipulates that participant location information is reported with a maximum precision of 0.1 degrees (equivalent to 11 x 11 km at the equator), that participants from grid locations with populations of less than 500 people are not indicated in published maps, and that there will be no public deposition of genomic sequences.

Additional approval for study of deidentified data from hospital records of hydrocephalus patients (postinfectious, non-postinfectious, spina bifida) since 2001 was granted by the CURE Children’s Hospital of Uganda Institutional Review Board.

#### 2.1.2 Clinical data

To investigate the relative risks (RR) of (i) PIH to NPIH and (ii) MM to NPIH, we fit two spatiotemporal statistical models to 19 years of clinical data collected by the CURE Children’s Hospital of Uganda (CCHU), a neurosurgical specialty care center with a nationwide catchment area. The clinical data included date of birth, date of first presentation at CCHU, village of residence and diagnosis (PIH/NPIH/MM) for each infant presenting with hydrocephalus from 2002 to 2020. The spatial coordinates are at a 1×1 km resolution while the temporal data are available at a daily resolution.

In our analysis, we excluded patients who were more than 90 days old at date of first presentation, as well as non-surgical patients, to minimize the risk of misdiagnosis between PIH and NPIH (and for consistency with MM). We approximate the locations of patients by calculating the population-weighted centroid coordinates for their village of residence, using the 100m gridded population data for 1-year-olds in Uganda obtained from WorldPop (*28*), averaged over the 19 years. We also exclude any infants from locations outside Uganda. These filters leave the respective numbers of individuals included in the study for each disease as n_*P IH*_ = 2061, n_*NP IH*_ = 856 and n_*MM*_ = 2199.

#### 2.1.3 Covariate data

With each individual infant, we associate the value of a range of covariates evaluated at the nearest location and time to their date and place of birth. Details on the resolutions of the datasets for each covariate, and how they were processed for compatibility with case data, are provided in Table S1. Genetic admixture and elevation are considered fixed over time. We incorporate genetic data in the form of a predicted surface constructed by fitting a geostatistical model to admixture data from a separate cohort (*4*) analyzed in (*29*). The only fine scale poverty data available was a snapshot from 2015, so we assumed that poverty distribution remained fixed over time. The available rainfall satellite remote sensing data vary daily over 0.1×0.1 degrees (11×11 km at the equator).

To investigate the mechanism behind the previously observed relationship between RR and rainfall, we consider different length trailing averages, ranging from 1 to 60 days. We also investigate the standardized precipitation evapotranspiration index (SPEI), which measures the extremeness of the rainfall for a given location, where negative values indicate drier-than-normal (drought) conditions and positive values indicate wetter-than-normal conditions. The details of SPEI computation are given in Section S1.2.

We also consider the EVI, a satellite product that illustrates the greenness of the surface of the earth (*30*) and improves upon the NDVI. Higher EVI values indicate greater vegetation vigor and photosynthetic activity, while lower values reflect sparse or stressed vegetation.

### 2.2 Modeling

#### 2.2.1 Overview and rationale

Our analysis uses geocoded residence locations and birth dates for infants with hydrocephalus ascertained through CCHU and classified into three hydrocephalus types (PIH, MM and NPIH; see Section 2.1.2). We treat the spatial locations and times of these observed cases as a realization of an underlying spatiotemporal point process.

A central challenge is that the intensity with which cases are ascertained at CCHU is influenced by access to care, referral pathways, and logistical constraints that vary over space and time. As a result, models that rely on an external population denominator may be biased if the observed cases do not constitute a complete or stable sample from the population at risk.

We therefore adopt a two-stage modeling strategy. First, we fit a population-referenced spatial bench-mark model for descriptive visualization only, to assess whether the spatial distribution of observed cases broadly follows that of the infant population. Second, for inference on associations with environmental and ancestry-related correlates, we use a case–control modeling framework with NPIH as an internal control group. Following the point-process formulation for case–control data described in (*31*), this approach conditions on observed case locations, eliminates the need for a population offset, and, under an explicit assumption about referral mechanisms, mitigates bias arising from unmeasured spatiotemporal variation in case ascertainment.

#### 2.2.2. Spatial benchmark model

As an initial descriptive check, we assess whether the spatial distribution of observed cases for each hydrocephalus type broadly follows the spatial distribution of the infant population, or whether there are systematic departures suggestive of additional spatial structure. Let λ_*d*_(**x**) denote the expected number of observed cases of hydrocephalus type d ∈ {PIH, MM, NPIH} per km^2^ at location **x** over the full study period. We fit the spatial benchmark model

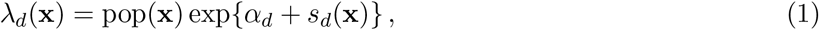

where pop(**x**) is the infant population density (*28*), α_*d*_ is a hydrocephalus-type-specific intercept, and s_*d*_(**x**) is a smooth spatial effect represented using thin plate regression splines. Details on spline fitting are provided in Supplementary Section S4.1. In this formulation, exp{s_*d*_(**x**)} captures departures from proportionality between the observed case intensity and infant population density, highlighting locations where case ascertainment is higher or lower than expected under simple population scaling.

Because external population surfaces include subpopulations that are not represented in the hospital referral data (notably within refugee settlements), and because case ascertainment at CCHU varies across space and time due to access and referral mechanisms, model (1) is used solely for exploratory visualization.

The fitted intensity surfaces are not interpreted as population-level incidence or absolute risk, but serve only as a descriptive spatial benchmark motivating the subsequent case–control analyses.

#### 2.2.3 Case–control spatiotemporal modeling framework

To study associations with environmental and ancestry-related covariates while avoiding reliance on an external population denominator, we adopt a case–control modeling framework with NPIH as an internal control group. This approach follows the point-process case–control argument in (*31*), in which conditioning on the observed event locations simplifies the model to a binary regression for case–control labels and removes the nuisance population intensity.

For each focal hydrocephalus type g ∈ {PIH, MM}, we define a binary outcome for the jth observed case at spatial location **x**_*j*_ and time t_*j*_:

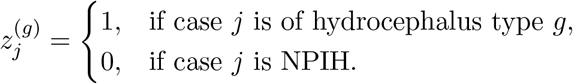

We assume

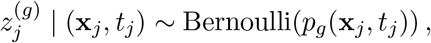

where p_*g*_(**x**, t) denotes the probability that an observed case at location **x** and time t is of hydrocephalus type g rather than NPIH.

##### Referral-bias assumption

This formulation estimates associations with hydrocephalus-type odds among infants whose cases were ascertained at CCHU, rather than attempting to estimate population-level incidence. It is robust to unmeasured spatiotemporal variation in case ascertainment under the assumption that unobserved factors influencing the probability of ascertainment act similarly on PIH/MM and NPIH after accounting for measured covariates and smooth spatiotemporal effects. Under this assumption, spatiotemporal variation in access to care, referral practices, or health system capacity is absorbed into the intercept and smooth terms, allowing covariate associations to be interpreted without bias from unknown referral intensity (*31*). This is particularly appropriate here because external population surfaces include substantial non-Ugandan subpopulations concentrated in refugee settlements (Figure S5) that are not represented in the hospital referral data, as medical care in these settings was provided through separate camp-based systems (see Supplementary Material).

#### 2.2.4 Spatiotemporal generalized additive model

For each focal hydrocephalus type g ∈ {PIH, MM}, we model the log-odds of type g versus NPIH using a spatiotemporal generalized additive model (GAM):

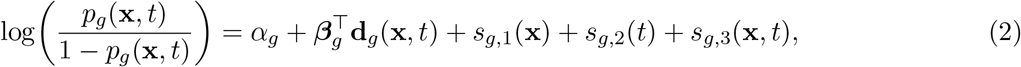

where **d**_*g*_(**x**, t) is a vector of covariates evaluated at location **x** and time t (including, where relevant, temporally lagged environmental summaries aligned to pregnancy windows), and ***β***_*g*_ are the associated regression coefficients. The smooth terms s_*g*,1_(**x**) and s_*g*,2_(t) capture residual spatial and temporal structure not explained by covariates.

Models were fitted using the mgcv package in R and all smooth terms are represented using spline bases within the GAM framework (*32*) as described in Supplementary Section S4.1. Spatial smooths are modeled using thin plate regression splines, and the space–time interaction, when included, is represented via a tensor-product smooth to accommodate differing spatial and temporal scales (see Supplementary Section S4.3.1). Smoothing parameters were estimated by restricted maximum likelihood (REML), which has been shown to provide stable smoothing parameter estimation and reduced overfitting compared with alternative criteria (*33*). While the smoothing parameter controls the effective complexity of a smooth term through penalization, the basis dimension determines its maximum allowable flexibility. Basis dimensions were set sufficiently large to ensure that the fit was not artificially constrained and were checked using the standard diagnostic tools, as illustrated in Supplementary Section S4.3.

#### 2.2.5 Model specification and selection

Model specification was guided by an exploratory inspection of marginal covariate relationships and by information-theoretic comparison of candidate models. For each focal hydrocephalus type g and each covariate, we first fitted restricted versions of Equation (2) including the covariates one at a time, along with smooth spatial and temporal effects (and, where supported, a space–time interaction), to assess plausible functional forms. Both linear and smooth representations of covariate effects, described in Equations (S2a) and (S2b) respectively, were examined and, where nonlinear terms were not supported, we proceeded with linear covariate effects.

For temporally varying covariates, we compared biologically plausible lag structures using AIC. For PIH, we evaluated trailing averages of rainfall over windows from 1 to 60 days before birth, motivated by evidence that relevant infections occur shortly after birth (*5, 9*). For MM, we evaluated monthly vegetation index (EVI) lagged between 0 and 10 months before birth to align with pregnancy windows.

Final multivariable models were selected by comparing candidate specifications using AIC. Biologically implausible combinations of covariates were excluded a priori (details in Supplementary Section S4.3). Model selection was based on information-theoretic criteria (AIC) rather than formal hypothesis testing; accordingly, no multiple-testing correction was applied, and model comparisons are interpreted as exploratory assessments of relative support rather than confirmatory inference.

Evidence for inclusion of the space–time interaction term s_*g*,3_(**x**, t) was assessed by comparing nested models with and without this term using a generalized likelihood ratio test. When the interaction was not supported, the additive model with s_*g*,3_ omitted was used.

## 3 Results

Section 3.1 presents exploratory analyses aimed at describing empirical spatiotemporal patterns and informing model specification. These steps include visual inspection of temporal and spatial trends, evaluation of marginal covariate relationships, and comparison of alternative lag structures using information criteria. Because these procedures are data-driven, they are interpreted as exploratory and hypothesis-generating. Confirmatory inference regarding covariate associations and space–time interaction is based solely on the prespecified multivariable case–control models reported in Section 3.2.

### 3.1 Descriptive spatiotemporal patterns

We begin by summarizing descriptive temporal, spatial, and covariate patterns in the observed hydrocephalus case data. These analyses provide contextual insight and motivate the subsequent spatiotemporal modeling, but are not interpreted as population-level incidence or absolute risk.

#### 3.1.1 Temporal patterns

Figure 1 summarizes temporal variation in the observed numbers of cases, scaled by infant population, for each hydrocephalus type. NPIH shows a fairly steady increase over the study period, consistent with changes in referral patterns and case ascertainment as awareness of specialized care expanded. In contrast, PIH and MM exhibit non-monotonic temporal patterns. Observed PIH cases increase rapidly before peaking around 2011 and then decline, whereas MM cases peak later, around 2018, following a modest decline between 2011 and 2015. Across all three hydrocephalus types, fewer cases are observed in 2020, coinciding with COVID-19–related travel restrictions.

**Figure 1.**
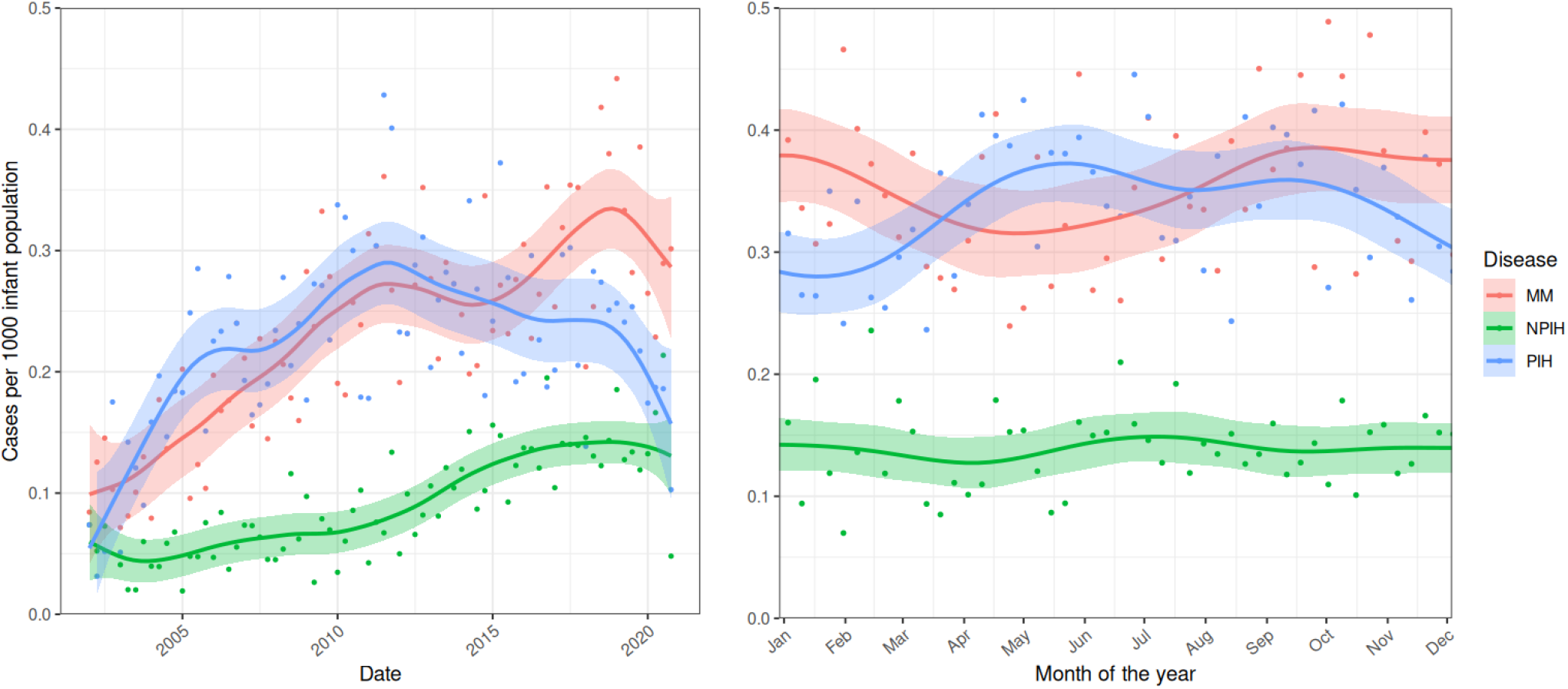
Observed cases per 1000 infant population aggregated by quarter (left) and by week of the year (right) for each hydrocephalus type.

The right panel of Figure 1 summarizes within-year variation in observed case counts to explore potential seasonality. PIH shows suggestive increases aligned with the biannual rainy seasons, which typically peak around April and October, whereas NPIH shows no clear within-year pattern. MM exhibits a relative decrease in observed cases between approximately March and July. These within-year summaries are descriptive; formal assessment of temporally varying environmental covariates is based on the spatiotemporal models described below.

#### 3.1.2 Spatial patterns

Figure 2 compares the spatial distribution of observed cases aggregated over the study period to the average infant population distribution. To ensure confidentiality, all locations are geomasked by jittering reported case locations (see Methods). NPIH broadly tracks the spatial distribution of the infant population, consistent with its role as an internal control group. In contrast, PIH exhibits pronounced spatial concentration, including notably higher observed case density northwest of Lake Kyoga and in areas south and west of the hospital between Lake Kyoga and the northern edge of Lake Victoria. MM displays a different spatial pattern, with less concentration northwest of Lake Kyoga and more diffuse distribution across eastern regions.

**Figure 2.**
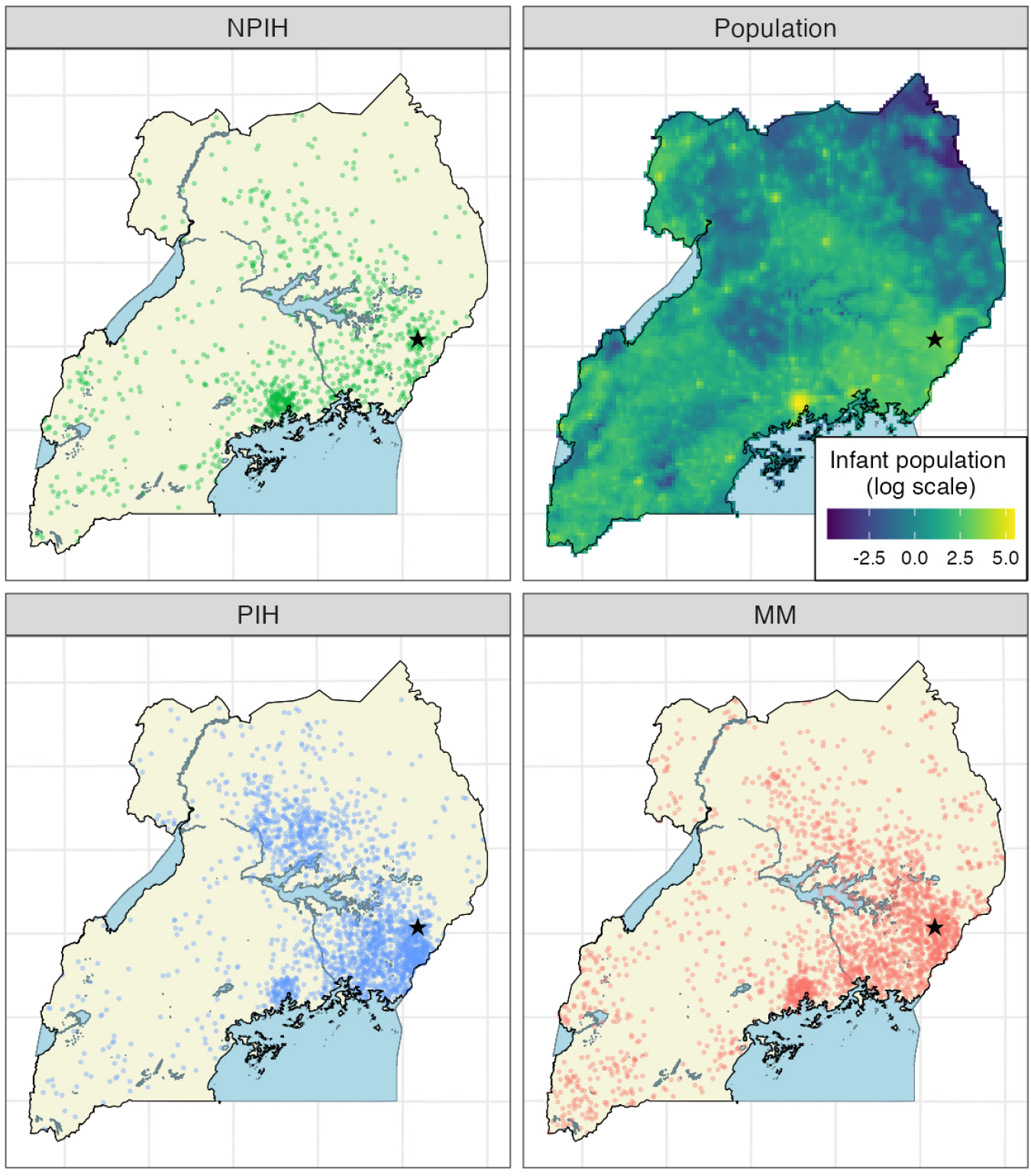
Spatial distribution of observed cases aggregated over the full study period for NPIH (upper left), PIH (lower left), and MM (lower right). Infant population density per km^2^ on a log scale is shown in the upper right panel. The location of CCHU is indicated by ⋆.

Figure 3 shows fitted spatial benchmark surfaces from Equation (1). After accounting for infant population density, NPIH shows minimal residual spatial variation relative to PIH and MM. In contrast, both PIH and MM exhibit clear spatial heterogeneity, with elevated fitted intensity for PIH north of Lake Kyoga and southwest of the hospital, and comparatively higher fitted intensity for MM in eastern Uganda. These benchmark surfaces are descriptive summaries of spatial variation in the intensity of observed cases relative to the infant population surface. They are intended to highlight departures from simple population proportionality in hospital-ascertained cases, rather than to estimate true population incidence.

**Figure 3.**
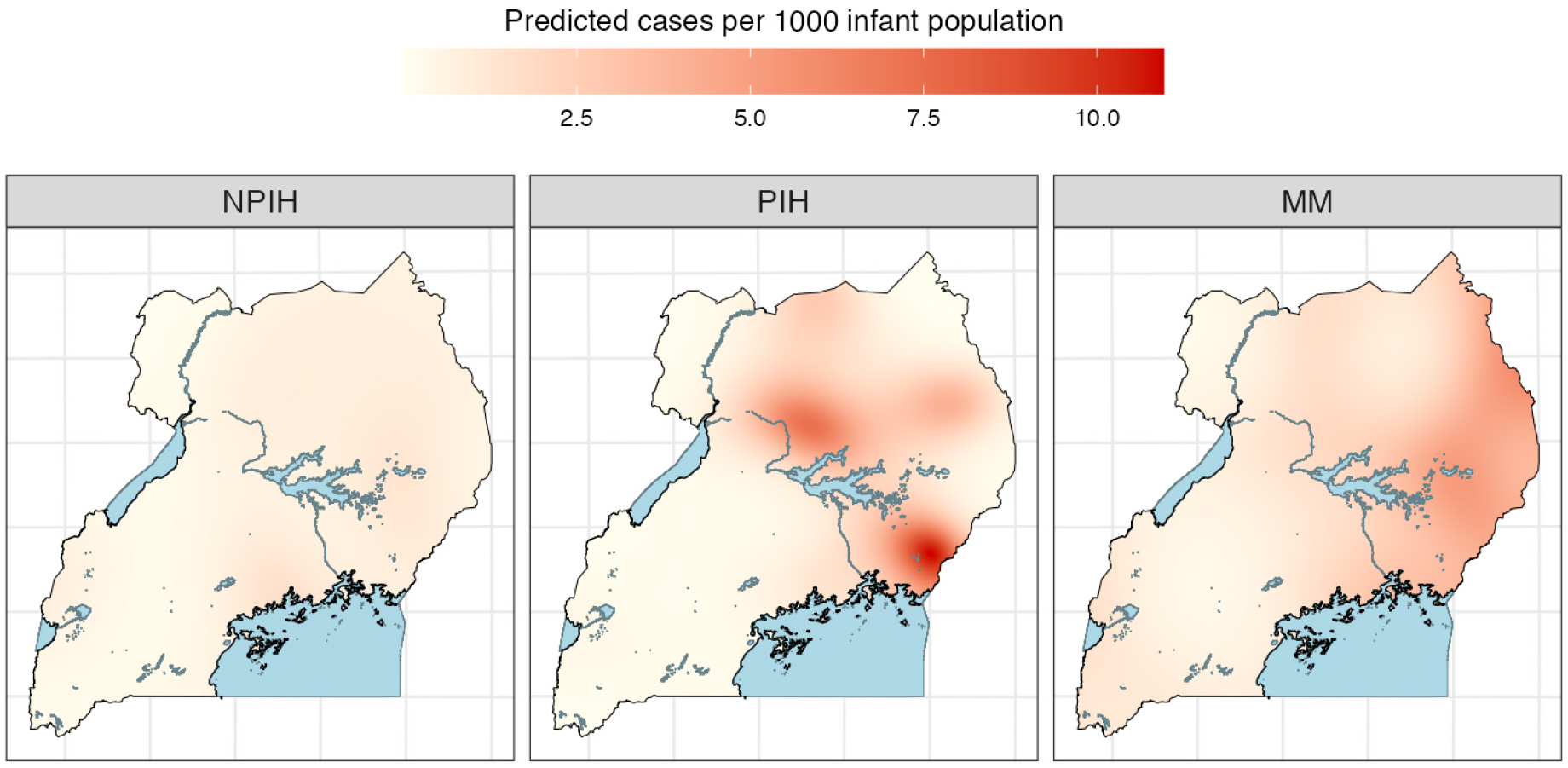
Spatial benchmark model predictions from Equation (1), shown as fitted cases per 1000 infant population per km^2^, averaged over the study period, for each hydrocephalus type.

#### 3.1.3 Exploratory covariate summaries and marginal associations

Figure 4 provides complementary descriptive summaries of the covariates considered in the analysis. The left-hand panels show the spatial variation of each covariate across Uganda, with temporally varying covariates averaged over the study period for visualization. The right-hand panels summarize modelbased marginal associations between each covariate and hydrocephalus-type odds relative to NPIH, after accounting for residual spatial and temporal structure.

**Figure 4.**
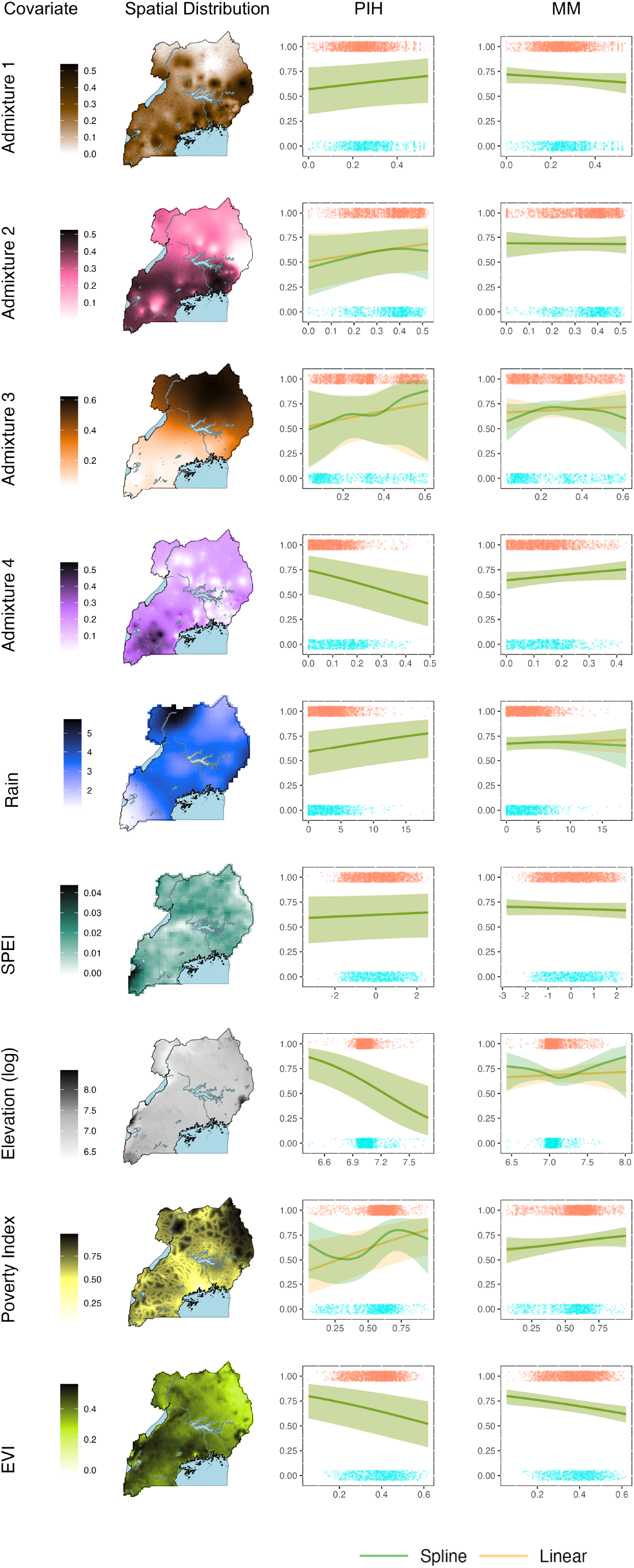
The left side plots depict the spatial variation for each covariate under consideration, with temporally varying covariates being averaged over time. For each disease, the plots on the right demon-strate the marginal relationships between each individual covariate (x axis) and the predicted RR computed assuming (a) a linear relationship (yellow), as in model (S2a) and (b) allowing for nonlinearity (green), as in model (S2b), after accounting for residual spatial and temporal effects. Estimates of the log-RR were constructed by averaging predictions 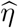 across a grid of spatiotemporal points for the full range of values of each covariate. Approximate 95% confidence bounds were derived similarly, by averaging 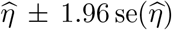 over the same grid. These estimates were transformed back to the RR scale via the inverse logit transformation. Red dots represent observed covariate values at the time and location of each observed PIH or MM case in the respective plots, and blue dots for NPIH cases. Units for the covariate values are in Table S1.

Across covariates, the estimated marginal associations were broadly consistent with linear effects over the observed covariate ranges. Although elevation, poverty, and genomic admixture showed hints of nonlinearity in places, uncertainty bands overlapped substantially, and these exploratory summaries did not provide strong evidence that nonlinear terms were required. Accordingly, linear covariate effects were used in the multivariable spatiotemporal models reported below.

#### 3.1.4 Temporal alignment of environmental covariates

For temporally varying environmental covariates, biologically motivated lag structures were examined to identify time windows most consistent with the observed data. Figure 5 summarizes these comparisons. For PIH, the best-supported models incorporated rainfall aggregated over the 14 days preceding birth, consistent with evidence that relevant neonatal infections occur shortly after birth. For MM, the best-supported models incorporated vegetation index (EVI) lagged by 8 months, corresponding approximately to early pregnancy for infants born with spina bifida.

**Figure 5.**
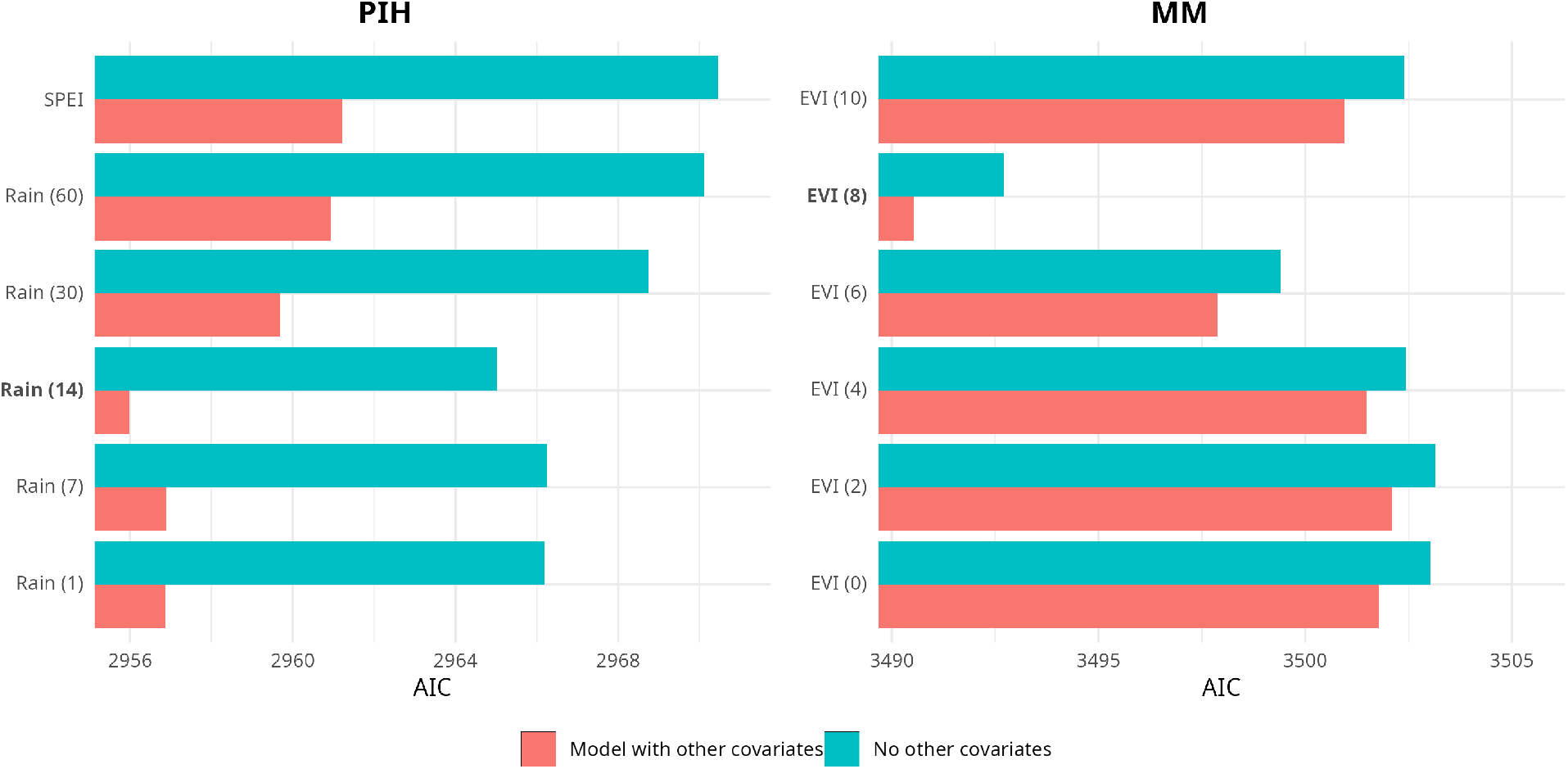
Comparison of lag structures for temporally varying environmental covariates. Labels Rain (*n*) correspond to trailing averages of *n* days prior to birth, and labels EVI (*k*) correspond to vegetation index lagged by *k* months. In bold with highlight the lags that minimise the AIC.

### 3.2 Spatiotemporal case–control models

#### 3.2.1 Selected covariates and space–time interaction

Based on information-theoretic model comparison (see Methods and Supplementary Material), a consistent set of covariates emerged, with strong support, for explaining variation in hydrocephalus-type odds relative to NPIH. For PIH, the final model included rainfall (14-day trailing average prior to birth), elevation, poverty, and genomic admixture 4. For MM, the final model included EVI lagged by 8 months and genomic admixture 4, with poverty providing a closely competing alternative specification.

Evidence for a space–time interaction was supported for PIH but not for MM. A generalized likelihood ratio test comparing models with and without the interaction term yielded p = 0.03 for PIH and p = 0.16 for MM. Accordingly, the interaction term was retained in the PIH model and omitted for MM.

#### 3.2.2 Estimated covariate associations

Figure 6 summarizes estimated covariate effects, expressed as log-odds ratios with 95% confidence intervals, from the final spatiotemporal models. For PIH, poverty and rainfall averaged over the 14 days preceding birth are positively associated with PIH odds relative to NPIH, whereas elevation and genomic admixture 4 are negatively associated. For MM, genomic admixture 4 is positively associated with MM odds, whereas EVI lagged by 8 months is negatively associated.

**Figure 6.**
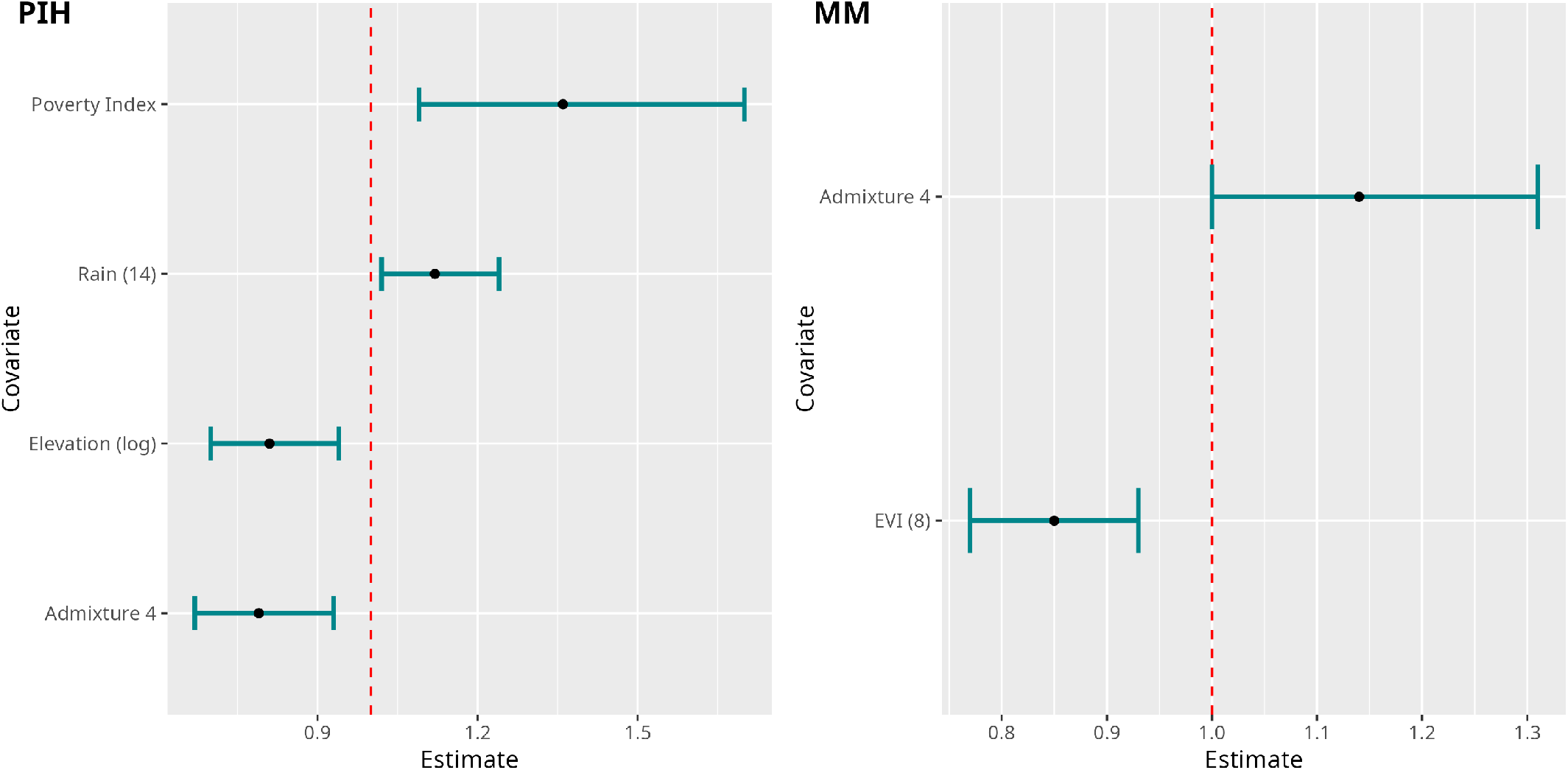
Estimated covariate effects (odds ratios) and 95% confidence intervals from the final spatiotemporal case–control models.

#### 3.2.3 Spatiotemporal exceedance probability maps

Figure 7 maps the exceedance probability (EP) that the modeled hydrocephalus-type odds for PIH and MM, each relative to NPIH, exceed their respective reference thresholds

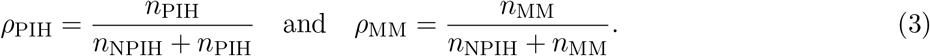

**Figure 7.**
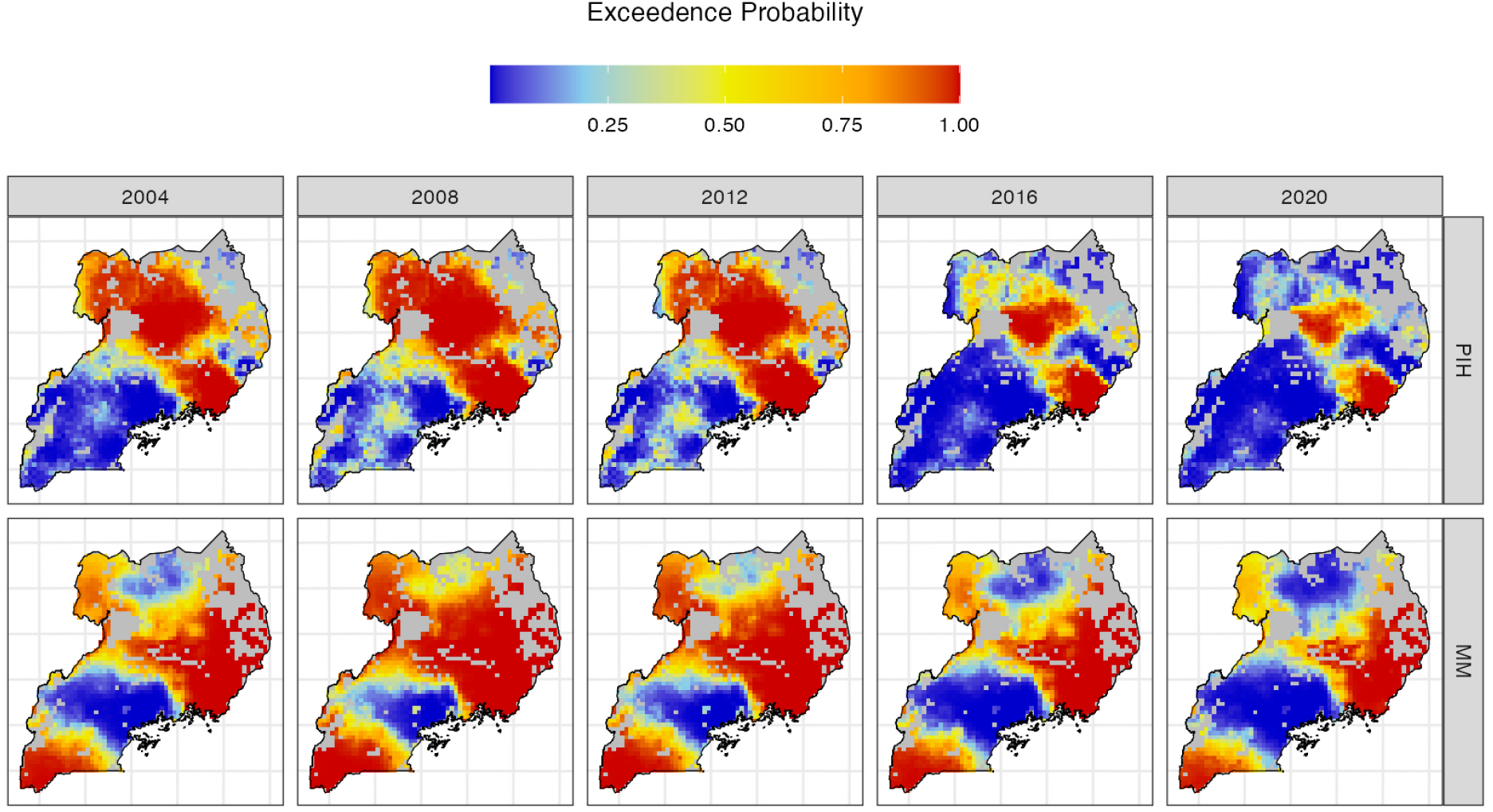
Exceedance probability maps showing the probability that hydrocephalus-type odds for PIH and MM, each relative to NPIH, exceed the respective reference thresholds ρ_PIH_ and ρ_MM_. Results are shown at four-year intervals (end date indicated), highlighting regions and periods with strongest evidence of elevated PIH or MM odds relative to NPIH.

These thresholds correspond to the overall proportions of PIH and MM among observed cases and provide a natural reference level for spatial comparison. Locations with higher EP indicate areas where the model provides strong evidence that hydrocephalus-type odds are elevated relative to this reference.

The EP maps provide finer spatial detail than the benchmark surfaces in Figure 3 and, shown at four-year intervals, illustrate changes over time. For PIH, elevated EP becomes progressively more concentrated north of Lake Kyoga and in east-central Uganda, including areas surrounding CCHU and extending south towards Lake Victoria. For MM, elevated EP is more geographically extensive, with lower EP in parts of the mid south-west and central-north regions. Over time, MM shows expansion of areas with high EP through approximately 2008–2012, followed by contraction towards more focused regions by 2020. Corresponding maps on the relative odds scale are provided in Figure S8.

## 4 Discussion

These findings demonstrate substantial evidence that prenatal environmental exposures predict the risk of hydrocephalus. Such effects modulate the risk associated with more static variables such as poverty, elevation, and genomic ancestral admixture at the population level. Our findings suggest that gene-environment interactions may underlie an important component of the risk of African infant hydrocephalus.

Although PIH has been noted to have an association with rainfall, it has never been examined at a fine spatial and temporal resolution, and the timing of rainfall has not been locally optimized to several weeks before birth. Such timing is consistent with what has been found for Melioidosis (*34*), where a similar 14 day window for rainfall has been shown to affect subsequent disease severity and mortality (*35*). The assumption for Melioidosis is that the bacteria found beneath the soil surface during dry seasons, emerge to colonize the surface as the rainy seasons begin (*36*). Consistent with this is our finding that lower elevations, where surface water will accumulate, are at increased risk. Similarly consistent is that more severe poverty in a setting such as Uganda, substantially increases the risk of environmental infection. That these infection risks from environmental exposures to infectious agents would be highly correlated with genomic ancestry at the population level was entirely unexpected. Nevertheless, such a gene-environment interaction is consistent with the growing appreciation of the relationship of genetic variation with infection susceptibility (*12, 13*).

These PIH findings were in stark contrast with MM risk. MM, a manifestation of spina bifida that frequently leads to hydrocephalus following surgical closure (*7*), is well known to occur upon a complex genetic background (*37*). It is largely preventable with adequate levels of dietary folate (*7*). An important source of dietary folate is green leafy vegetables (*38*). The protective association of EVI and reduced risk of MM was very substantial. Also notable, was that the same ancestral genomic admixture that was protective for PIH, appeared to significantly elevate the risk of MM in this study. It could be that common variants in the haplotypes that define those admixtures contain gene variations that impact both the immune system as well as neural tube development. As both PIH and MM have their own specific geographical distribution in Uganda, it could also be that environmental risks that contribute to PIH and MM are spatially enriched in regions that have high proportions of specific genomic admixtures. These results provide substantial evidence for associations between gene-environment interactions and infant hydrocephalus, which have not been previously reported. Both the PIH and MM associations are consistent with our understanding of the biology and ecology of these diseases. That the associations are all observed prenatally suggests that timing is a critical factor to more effectively predict and prevent such conditions.

There are important limitations for these findings; paramount is that association does not prove causation. Follow-on studies are required to validate and replicate these spatial and temporal results, and to prove that public health measures built upon them can indeed reduce the risk of these devastating conditions. An additional limitation is that a single referral site dominated the treatment of hydrocephalus in Uganda during these 19 years. We controlled for such referral bias through the use of NPIH cases as controls. As discussed in Section S3, NPIH is an ideal control for PIH because the signs of disease that motivate referral, an enlarging head in an infant, are identical. Although CCHU has also been the dominant site for MM referral during the study, the different clinical presentation (open spine) means that NPIH is not an ideal control for MM. Nevertheless, any referral bias due to distance from the hospital should be similar for NPIH and MM, and it should also help control for alternative sources of medical care for infants within refugee camps that are absent from our case data. Lastly, the societal lockdown put in place in March of 2020 substantially affected travel within the country, and although we note in Figure 1 that the decreased case referrals during this time was rather uniform across each case type, it is not clear whether the impact of these restrictions would be the same for all diseases.

## 5 Conclusion

PIH and MM contribute the majority of infant hydrocephalus in Uganda. The observed geographical clustering and association of PIH with poverty and rainfall point to the importance of localizing the source of the most common bacteria leading to PIH in this country - highly pathogenic strains of *Paenibacillus thiaminolyticus* (*3–5*). A combined approach, localizing village environmental sources and initiating public health policies to reduce the pathways of infection due to regional newborn care practices, is advisable. The association of MM risk with vegetation, and presumably crop health, suggests that strategies might focus folic acid fortification and supplementation on the regions and times with highest risk. Whether agricultural planting strategies, in a region where subsistence farming dominates the rural population, might be an additional strategy to lower MM risk is an intriguing possibility raised by these findings. Because some of the genetic association with MM is related to folic acid metabolism gene variations (*39*), our findings of a population stratification by ancestral admixture, preserved geographically, may indicate that optimizing the recommended dose of folic acid may be required across those populations.

## Data Availability

Due to patient confidentiality restrictions, individual-level birth date and geographic location data used in this study cannot be publicly released. To enable reproducibility of the analysis pipeline, we provide a surrogate dataset in which these personal identifiers have been progressively randomized. This surrogate dataset, together with the full analysis code, reproduces the modeling workflow. The surrogate data and code are available in the project repository. Access to the original data may be considered upon reasonable request and subject to institutional ethical approvals.

## 6 Acknowledgements

This work was supported by an NIH Director’s Transformative Award R01AI145057 (SJS).

## 7 Author Contributions

SJS, JK and CF conceived and designed the project. BKN, DN, MW, PS, and EM-K, performed the data organization and verification. JM, EN, BCW, PS, JN Performed the clinical care. LN, RM, MS, MM, JNP, PJD, ANW, CF and SJS Performed the statistical analysis. SUM and MM performed the ancestral genomic analysis. LN, PS, EM-K, JEE, JNP, CF and SJS Contributed to the writing of the article. CF, LN, PJD, and SJS reviewed the manuscript and analysis and did the final editing. All authors approved the final manuscript.

