## Supplementary Material for "Spatiotemporal patterns of African infant hydrocephalus are predicted by prenatal environment and ancestral genomics"

March 10, 2026

### S1 Covariate data

#### S1.1 Sources

Table S1 provides information on the covariate data used in our modeling process. The 'Description' column gives details about the original version of the dataset and 'Source' provides a link and reference to it. 'Modifications' describes the modifications we made to render the original data compatible with our case data, including projection to different coordinate reference systems and spatial and temporal aggregation/disaggregation.

#### S1.2 SPEI

The SPEI data were generated from the existing temperature and rainfall datasets detailed in Table S1. Specifically, for each location $\mathbf{x}$, the SPEI at time $t$ was calculated by subtracting the potential evapotranspiration from the total precipitation for a time window of 3 months centered about $(\mathbf{x}, t)$ and comparing this short-term difference to the long-term average difference. The SPEI is the number of standard deviations the short-term value is from the long-term average, where positive values indicate higher than average precipitation and vice versa. To compute the potential evapotranspiration we used the Hargreaves method [1] amended using the recommendation due to Droogers and Allen [2], which corrects for the amount of rain each month as a proxy for insolation. The mean external solar radiation required for the computation was estimated from the latitude and month of the year. The SPEI was then derived using the `SPEI` function in `R` [3].

#### S1.3 Genetic Admixture

In [4], we used the ADMIXTURE software to identify four genetic admixture groups within the cohort, for which each individual's genetic data and village of residence were available. These groups were referred to as admixture 1 to admixture 4. For each participant, the proportion of each admixture group was calculated such that the sum of all proportions equaled 1. We observed a clear spatial relationship between an individual's genetic ancestry and the region where they lived. By fitting four separate geostatistical models to these data, we were able to predict the distribution of the proportion of each admixture across the country. Those predicted surfaces are shown on the left side of Fig. 4 and these are the values used as covariates in the model for this present paper.

Table S1: Description of the covariate data used in modeling PIH and MM relative risk across Uganda.

| **Covariate** | **Description** | **Resolution (spatial/ temporal)** | **Source** | **Modifications** |
|---|---|---|---|---|
| Population of Uganda | Unconstrained [5] estimates of total number of people aged < 1 year per grid square broken down by gender using the mapping approach of [6]. | 0.000833°≈ 100m / Yearly | WorldPop [7] | We used the unconstrained version because it was available yearly throughout the study period.<br>Infant population derived by summing males and females.<br>Coordinates reprojected from WGS84 (° lat/long) to UTM Zone 36N (meters).<br>Monthly temporal resolution approximated via linear interpolation. |
| Temperature: tmin/tmax | Maximum and minimum daily temperatures in °C recorded at weather stations across Uganda averaged for a given month and interpolated to a grid. | 0.5°≈55km / Monthly | CRU TS 4.06 [8] | We disaggregated the spatial resolution to 0.1°, to agree with the RFE2 rainfall data for use in computing SPEI. |
| Poverty rate | Proportion of people (0-1) per grid square of Uganda living on less than $1.25 a day in 2011. | 0.00833° ≈1km / NA | WorldPop (2011) | Aggregated (mean) to 0.1° grid resolution.<br>Coordinates reprojected from WGS84 (° lat/long) to UTM Zone 36N (meters). |
| Enhanced vegetation index (EVI) | Estimates vegetation coverage by analyzing satellite data. Observed values range from -0.2 to 1 with increasing values corresponding to increasingly dense vegetation. Negative values indicate urban areas/water, values below 0.2 indicate very sparse vegetation. | 0.05°≈5.5km / Monthly | MODIS: Terra Vegetation Indices (MOD13C2) [9] | Coordinates reprojected from WGS84 (° lat/long) to UTM Zone 36N (meters). |
| Rainfall (RFE2) | Total rainfall in mm/day calculated by combining GTS rain gauge data, AMSU microwave, SSM/I satellite, and GPI IR estimates. | 0.1°≈11km / Daily | NOAA FEWS RFE2 | Trailing averages per grid square computed for window lengths ranging from 7-60 days.<br>Monthly averages computed for use in SPEI computations.<br>Coordinates reprojected from WGS84 (° lat/long) to UTM Zone 36N (meters). |
| Admixture | Predicted proportion (0-1) of Admixtures 1-4 making up the DNA of individuals residing at each grid square. Computed using a geospatial model fit to genetic data [4]. | 0.1°≈11km / NA | — | Computed using geostatistical models as described in Section S1.3. |
| Standardised precipitation evapotranspiration index (SPEI) | Compares recent rainfall to long-term average considering evapo-transpiration, for drought/ wetness detection. Units are the number of standard deviations the observations are from the long term mean. | 0.05°≈5.5km / Monthly | — | Computed using rainfall and temperature data as described in Section S1.2. |

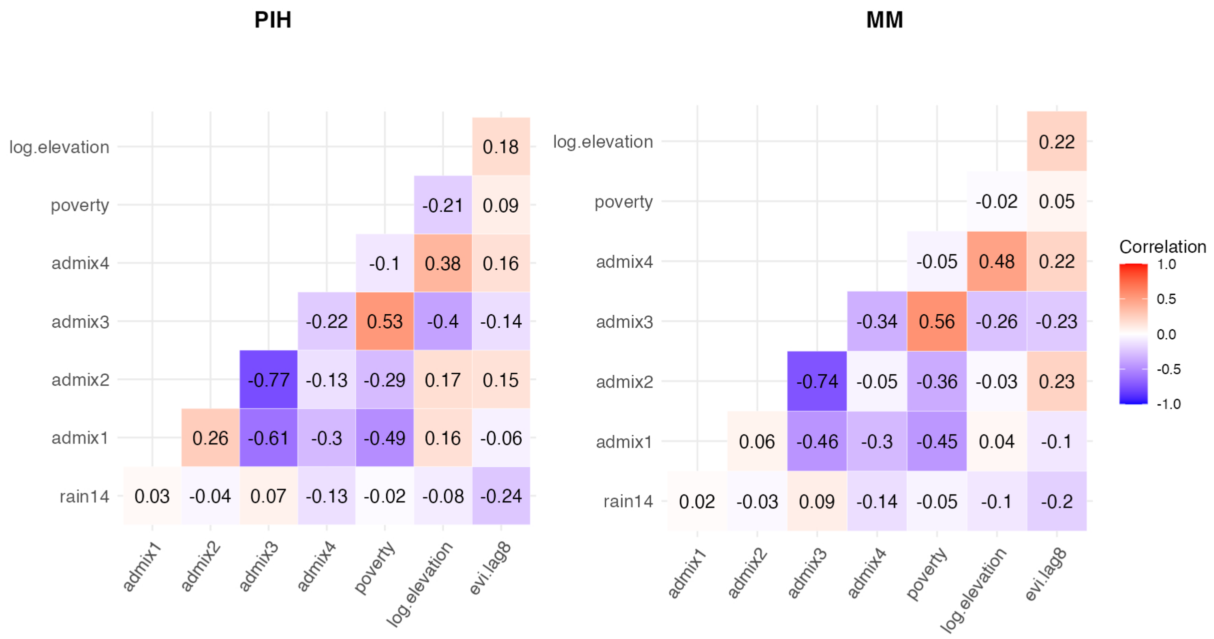


Figure S1: Correlations between covariates evaluated at case observations for PIH (left) and MM (right).

#### S1.4 Covariate relationships

Figure S1 shows the correlations between different covariates evaluated at the locations of the respective cases. We observe highest correlations between the admixture surfaces for both diseases, particularly between admixture 2 and admixture 3. This co-linearity paired with the dependency enforced by the construction of the surfaces (which must sum to 1, see Section S1.3) means we remove models with multiple admixture variables from consideration.

Figure S2 shows the cross correlation between rainfall and EVI. There are biannual rainy seasons in the East African Highlands. We see that there is a clear seasonal relationship with EVI correlated with rainfall one month prior, but also with rainfall 7 months prior, 5 months after, 11 months after, etc. Figure S3 shows that the lag with the greatest correlation is sometimes one of these time points. This explains why our exhaustive search for covariate selection in our PIH model sometimes chooses EVI, lagged by 6-8 months, over rainfall, lagged by 14 days. Given that there is no biological reason that EVI from 8 months prior to birth would affect infection risk in PIH cases after birth, we opt for the model that includes rainfall instead for PIH.

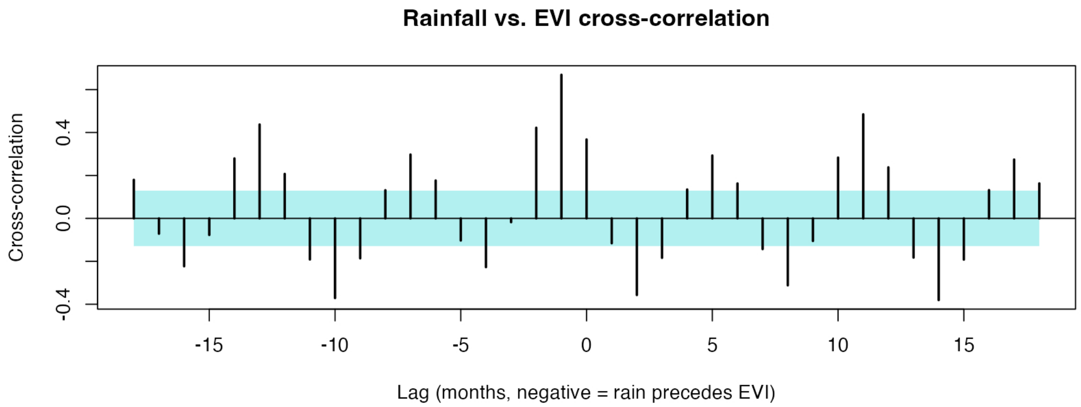


Figure S2: Cross correlation between Rainfall and EVI.

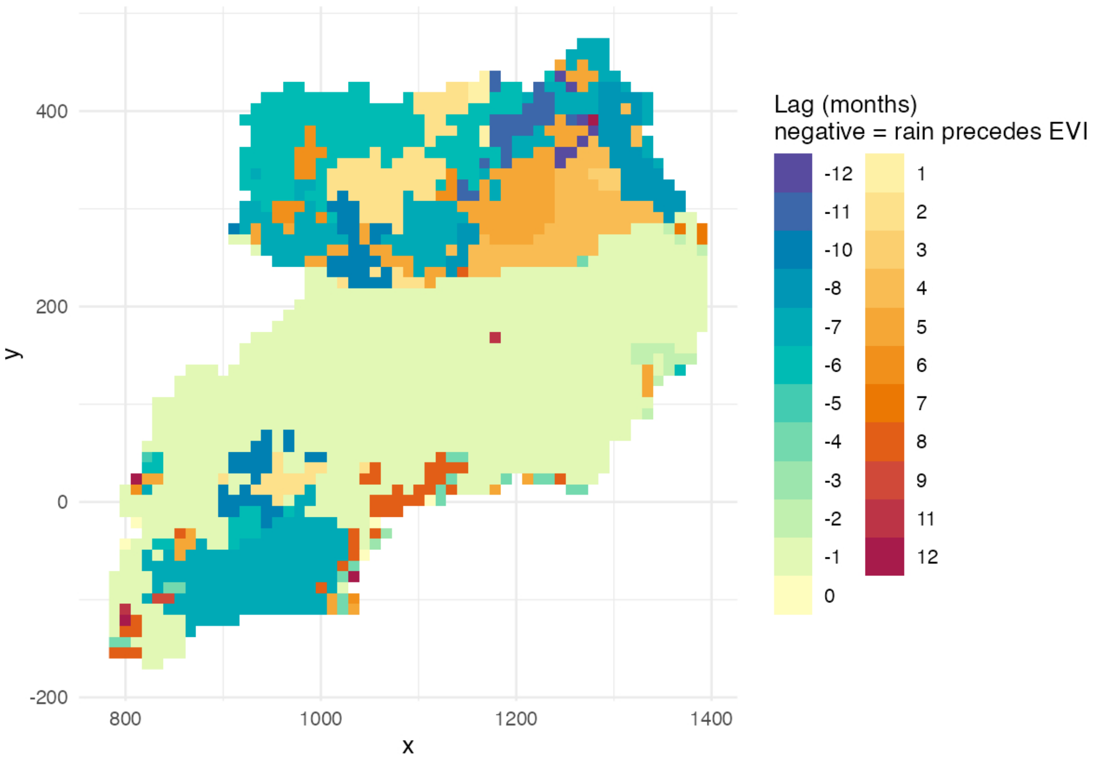


Figure S3: Cross correlation spatial variation.

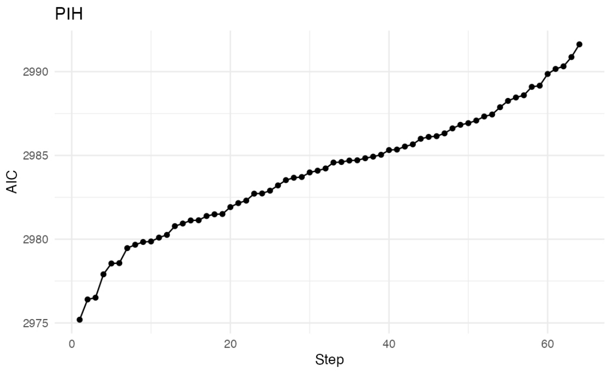


(a)

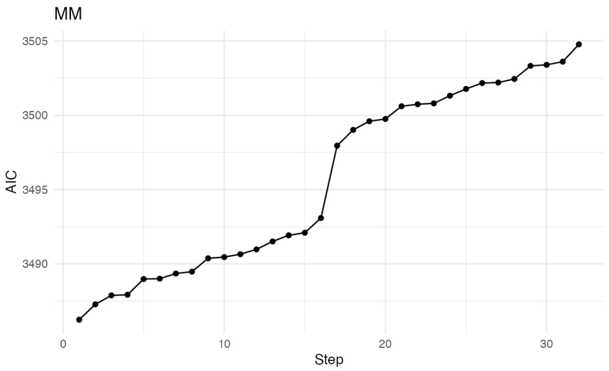


(b)

| PIH | | |
|---|---|---|
| Covariates | AIC | delta |
| admix4, rain14, log.elevation, poverty | 2975.19 | 0.00 |
| admix3, admix4, rain14, log.elevation, poverty | 2976.40 | 1.21 |
| admix2, admix4, rain14, log.elevation, poverty | 2976.51 | 1.32 |
| admix2, admix3, admix4, rain14, log.elevation, poverty | 2977.90 | 2.71 |
| admix4, log.elevation, poverty | 2978.55 | 3.36 |
| admix4, rain14, poverty | 2978.57 | 3.38 |
| admix3, admix4, rain14, poverty | 2979.47 | 4.27 |
| admix3, admix4, log.elevation, poverty | 2979.67 | 4.48 |

(c)

| MM | | |
|---|---|---|
| Covariates | AIC | delta |
| admix4, evi.lag8 | 3486.24 | 0.00 |
| admix3, admix4, evi.lag8 | 3487.27 | 1.03 |
| admix2, admix4, evi.lag8 | 3487.88 | 1.64 |
| admix4, evi.lag8, poverty | 3487.92 | 1.68 |
| admix3, admix4, evi.lag8, poverty | 3488.98 | 2.74 |
| admix2, admix3, admix4, evi.lag8 | 3489.00 | 2.76 |
| evi.lag8 | 3489.35 | 3.11 |
| admix2, admix4, evi.lag8, poverty | 3489.48 | 3.24 |

(d)

Figure S4: **Exhaustive selection by AIC:** Figures S4a and S4b plot the AIC of each of the models fit by exhausting all possible combinations of covariates, for PIH and MM respectively. The models are ranked from lowest AIC (best) to highest (worst) AIC, with step indicating the rank number. In Tables S4c and S4d, row number corresponds to step. Each row represents a model with a the given combination of covariates, fit according to 2. The best fitting models for each disease are in the top rows of the tables and have the lowest AIC. Subsequent models have AIC which is delta greater than the best model. Models with delta $\leq 2$ relative to the best model are considered statistically indistinguishable in terms of AIC and comparably supported by the data, and are outlined in green. Any model that includes two or more correlated covariates was eliminated.

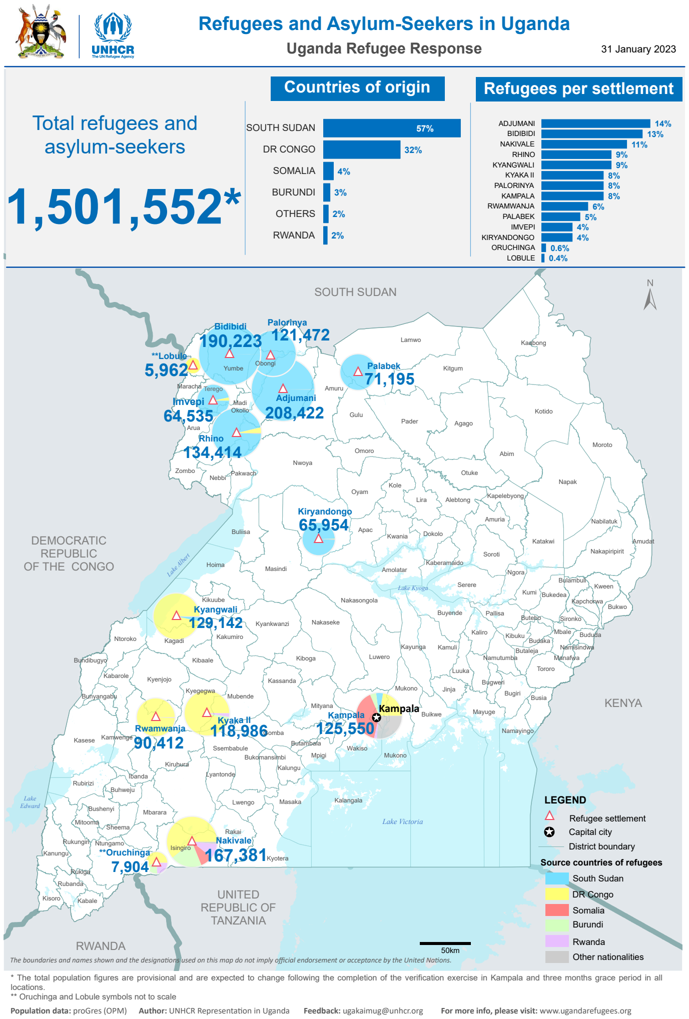


Figure S5: Locations and population of camps with refugees and asylum seekers. Figure reproduced under Creative Commons Attribution 4.0 International Public License, from the Operational Data Portal from the UNHCR at `https://data.unhcr.org/en/documents/details/98683`.

### S2 Model selection

Further results of exhaustive model selection are shown in Figure S4. We list the top 8 models ordered by AIC for PIH and MM. Note that including poverty as an additional covariate for MM produces several plausible models (with AIC comparable to the optimal model). Each of these more optimal covariates appear consistently in the better models. Although in the main paper we carry out our predictions for the optimal model, any plausible model performs similarly well.

### S3 Refugee camps

Figure S5 shows the locations of refugee camps, which are populated by non-Ugandan individuals who were not included within our hydrocephalus case dataset.

PIH and NPIH cases are indistinguishable prior to surgery, as both present with macrophaly. Neonates with either condition are referred to the CURE Children's Hospital of Uganda (CCHU) in Mbale. It follows that any referral biases should be the same for each disease and this forms the rationale to consider the relative risk (RR) of PIH to NPIH.

In contrast to hydrocephalic infants who present with an enlarging head, infants with MM are born with an open spine and a typically small head, only enlarging after the spine is surgically closed. Initially, we modeled the absolute risk of MM and controlled for population, rather than considering the relative risk of MM to NPIH. However, after comparing the spatial patterns of the population data with our case data, we discovered a discrepancy in the northwest and central-west regions, corresponding to the locations of several large refugee camps (Figure S5). The populations of these camps are incorporated into the population data set, but the residents were not included in our disease database, which meant that controlling for the population introduced selection bias. Therefore, we modeled the RR of MM to NPIH, as we did for PIH, to reduce this bias from our findings.

### S4 Model method

A generalized additive model (GAM) is a type of regression model that extends the class of generalized linear models (GLMs) to allow relationships between predictors and the outcome to be nonlinear by representing them as smooth functions. These smooth functions are typically constructed using basis expansions known as *smoothing splines*. Model (2) was fitted in R using the `mgcv` package, which implements GAMs using penalized regression splines and automatic smoothing parameter selection [10].

#### S4.1 Smoothing Splines

A smoothing spline represents an unknown function $s(\mathbf{x}_1, \mathbf{x}_2 ...)$ as a weighted sum of basis functions, which are defined relative to a set of knots. The choice and number of basis functions and knots influence the flexibility of the spline: too few can under-fit the data, while too many can lead to overfitting.

The actual complexity of the smoothing spline is determined by the smoothing penalty $\phi$, which penalizes the function's roughness by shrinking its coefficients. That is, the smoothing spline is chosen to minimize the penalized log-likelihood of the model

$$\mathcal{L}(\alpha, \boldsymbol{\beta}, s(k)) - \phi J(s(k)), \tag{S1}$$

where $\mathcal{L}$ is the log-likelihood, and $J(s)$ is a function quantifying the roughness of $s$. While the $k$ largest eigenvalues of the spline provide an upper bound on the effective degrees of freedom (EDF), $\phi$ determines how much of that flexibility is actually realized.

A key advantage of thin plate regression (TPR) splines over alternatives such as cubic regression splines or penalized splines is that they do not require the analyst to choose knot locations, which can influence the fit. In the full thin plate (TP) spline formulation, every data point $x_{i,j}$ acts as a knot, which yields a basis that is theoretically optimal but computationally impractical. TPR splines overcome this by retaining only the basis functions corresponding to the $k$ largest eigenvalues of the TP spline penalty matrix. This construction preserves the optimality of TP splines while avoiding the knot placement problem, making TPR splines well-suited for spatiotemporal models.

#### S4.2 Exploratory models

To examine the marginal relationships between each covariate under consideration we fit two models for each disease-covariate combination $(g, i)$. Model (S2a) forces any relationship between covariate $i$ and the log-odds of hydrocephalus type $g \in \{\text{PIH}, \text{MM}\}$ versus NPIH to be linear, while (S2b) allows for nonlinearity.

$$\log\left(\frac{p_g(\mathbf{x}, t)}{1 - p_g(\mathbf{x}, t)}\right) = \alpha^a_{g,i} + \beta_{g,i} d_{g,i}(\mathbf{x}, t) + s^{a_1}_{g,i}(\mathbf{x}) + s^{a_2}_{g,i}(t) + s^{a_3}_{g,i}(\mathbf{x}, t), \tag{S2a}$$

$$\log\left(\frac{p(\mathbf{x}, t)}{1 - p(\mathbf{x}, t)}\right) = \alpha^b_{g,i} + s^b_{g,i}(d_{g,i}(\mathbf{x}, t)) + s^{b_1}_{g,i}(\mathbf{x}) + s^{b_2}_{g,i}(t) + s^{b_3}_{g,i}(\mathbf{x}, t). \tag{S2b}$$

Here, $\mathbf{d}_{g,i}(\mathbf{x}, t)$ is a single covariate evaluated at location $\mathbf{x}$ and time $t$. The scalars $\beta_{g,i}$ are the associated regression coefficients for the linear relationships in (S2a), while the splines $s^b_{g,i}$ provides the non-linearlity

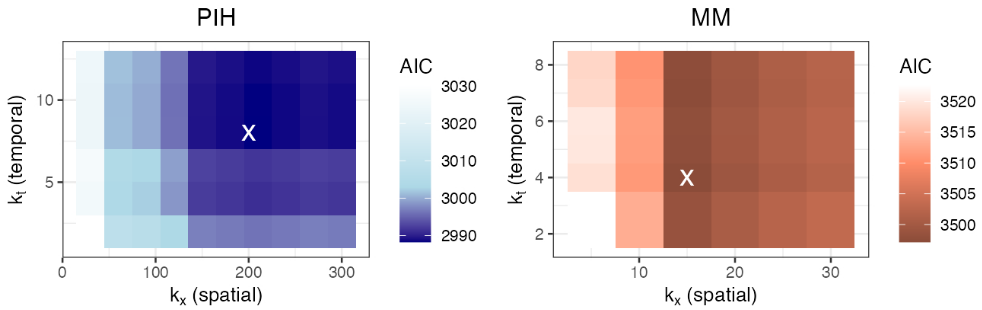


Figure S6: Optimization of $k_x$ and $k_t$, the numbers of basis functions for smooth functions modeling the main effects $s_x$ and $s_t$ in model (2), respectively. When these values are too low, the EDF saturates, and a higher than optimal AIC is observed. The lowest AICs for each disease occur at the values indicated with a white X.

in (S2b), and are fit as described in Section S4. Otherwise, the equation terms correspond to those in our main model (2), that is, the remaining smooth terms capture residual spatial and temporal structure and the optional space–time term captures any significant interaction.

#### S4.3 Optimization

To assess the quality of the fit for models formulated according to Equation (2), we used the Aikake Information Criteria (AIC). In the context of GAMs, this criterion is most appropriate, because unlike the Bayesian Information Criterion (BIC), it can be adjusted to account for the dependence of the EDF on the smoothing parameters estimated from the data. Alternatives such as ANOVA hypothesis testing, require models to be nested and may not be valid for penalized smooth terms [11, 10].

We compared two smoothing parameter estimation methods available in the `mgcv` package: restricted maximum likelihood (REML) and generalized cross-validation (GCV). REML is generally preferred because, while GCV is computationally efficient, it tends to under-smooth. In contrast, REML provides more stable smoothing parameter estimates which leads to more robust performance for inference, especially in non-Gaussian models with complex smooth structures such as spatial, temporal, or interaction terms [10]. This was borne out in our results; the AIC stabilized under REML, while GCV continued to increase the EDF without significant AIC improvement.

Additionally, if the number of basis functions is set too low, the EDF will saturate, underfitting the data and resulting in a higher than optimal AIC. This occurred for our final model (2), in which we have two smoothing splines for the main spatial and temporal effects, and a possible third if the interaction term is included. We refer to the numbers of basis functions for smooth functions modeling the main effects $s_x$ and $s_t$, as $k_x$ and $k_t$ respectively. Figure S6 shows the results of simultaneously optimizing these values for each disease. The AIC stabilizes at approximately $k_x = 200$ and $k_t = 8$ for PIH and $k_x = 15$ and $k_t = 4$ for MM. These are the numbers of basis functions we specify in our final models.

##### S4.3.1 Interaction term

Because the spatial and temporal variables have different units (kilometers versus months) and thus vary on different scales, a standard TPR spline cannot model the joint effect accurately in the interaction term. Instead, a tensor product smoother is used, as it allows the spatial and temporal dimensions to have separate smoothness parameters and scaling. Since our model already includes the main effects of space and time, the interaction is included using a tensor *interaction* term, which isolates only the joint variation beyond the additive effects and avoids overlap with the main effects [10]. This tensor interaction requires that two separate values, $k_{x'}$ and $k_{t'}$, are specified for space and time because the complexity of the smoothing in each dimension is controlled independently. To identify an appropriate level of complexity for the tensor product interaction term, model (2) was fitted, with the already optimized values for $k_x$ and $k_t$, for each of a grid of basis dimensions ($k_{x'}$, $k_{t'}$). We restricted our model selection to include only models with a stable

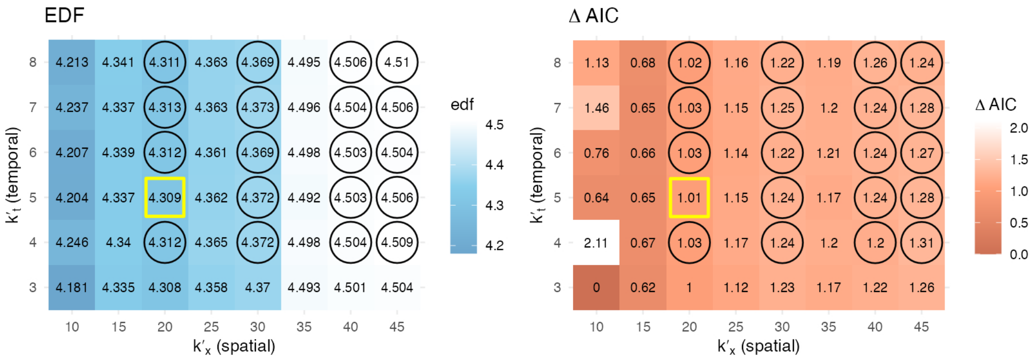


Figure S7: We optimized the basis dimensions for the tensor product interaction term by refitting model (2) for a grid of values of $k_{x'}$ and $k_{t'}$). The models included the previously optimized values for PIH: $k_x = 200$ and $k_t = 8$. We recorded the EDF and AIC, which vary irregularly as $k_{x'}$ and $k_{t'}$ increase, due to the simultaneous re-estimation of multiple smoothing parameters that occurs in an interaction smoothing spline. Each model's *stability* was assessed by examining the relative change in EDF with respect to increases in either $k_{x'}$ or $k_{t'}$. Models where the EDF changed by less than 1% per unit increase in either basis dimension were considered *stable* and are circled in black. Among these stable models, the one with the lowest AIC was selected as the optimal configuration, highlighted in yellow. For legibility we report the difference between the AIC and the minimum observed AIC for each model, referred to as Δ AIC.

EDF, which resulted in optimal values $k_{x'} = 20$ and $k_{t'} = 5$ as shown in Figure S7. Only results for PIH are included since the interaction term was not found to be statistically significant for MM.

#### S4.4 Predictions

Figure S8 shows the relative risk predicted at 4 year intervals over the study period across Uganda using model (2), corresponding to the exceedance probability maps given in Figure 7. The thresholds used as the breaking point (white) in the color scale are defined in (3). We also demonstrate the difference observed in prediction when comparing the models with and without the spatiotemporal interaction term in Figure S9.

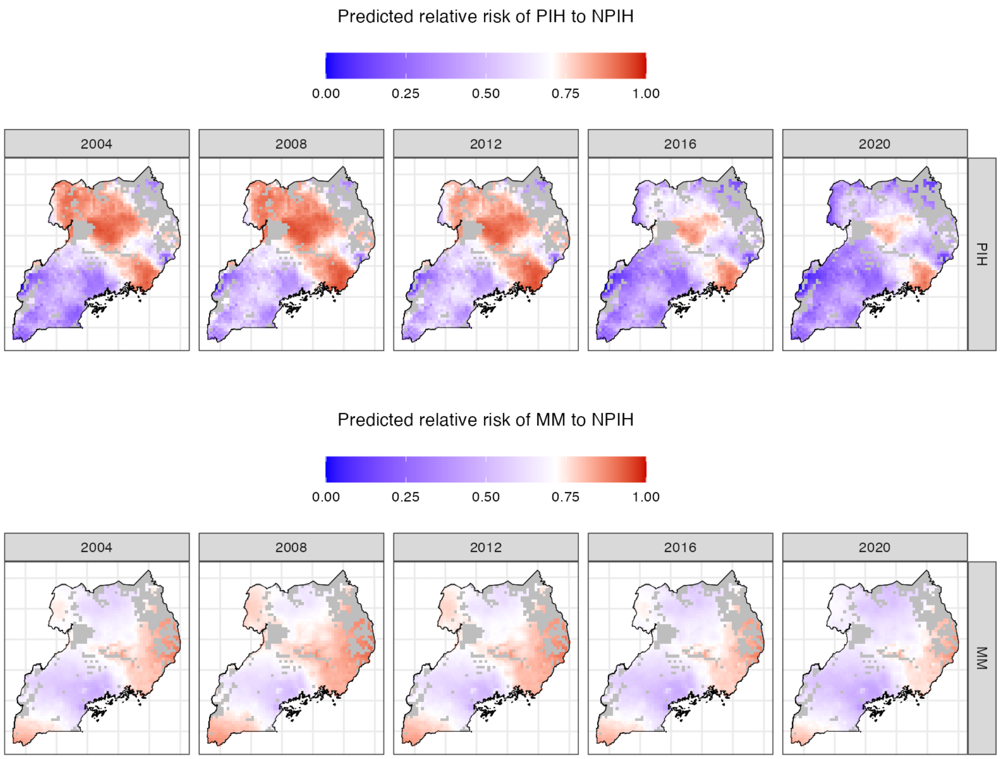


Figure S8: Model predicted relative risk (RR) over study period for each disease for four year intervals (end date indicated).

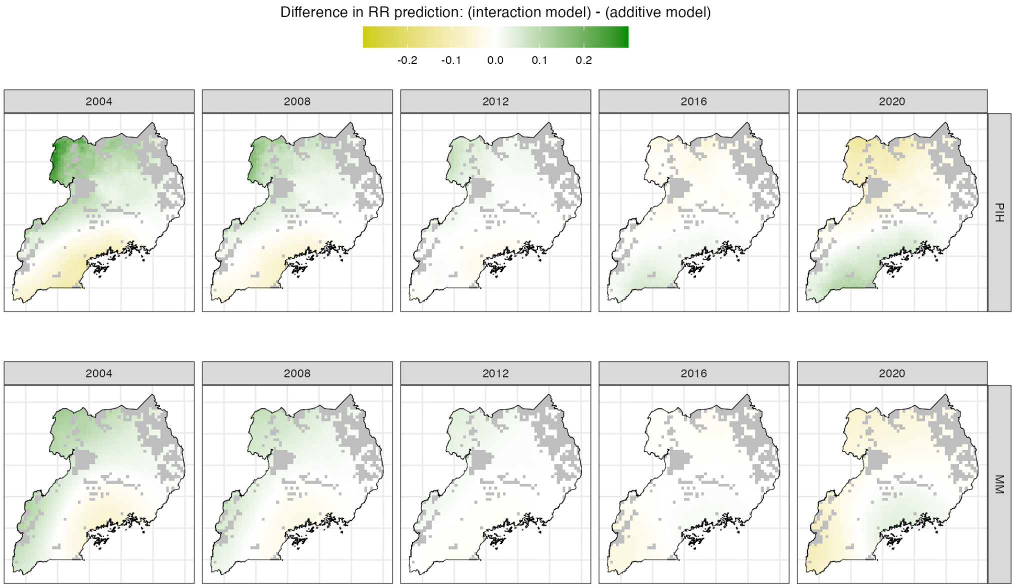


Figure S9: Comparing models with and without the spatial interaction term for four year intervals (end date indicated).
